# The Use of Psychoactive Substances in the Context of the Covid-19 Pandemic in Brazil

**DOI:** 10.1101/2020.09.25.20194431

**Authors:** César Augusto Trinta Weber, Ingridi Teixeira Monteiro, Julia Medeiros Gehrke, Wagner Silva de Souza

## Abstract

**Background:** The protocols for mitigating the transmission of Covid-19 seem to be a trigger for the use of psychoactive substances, as an individual’s adaptive response to support this new way of being in the world. Objective: To investigate the use of psychoactive substances in the context of the Covid-19 pandemic in Brazil. Method: A cross-sectional and prospective observational study, undertaken in a virtual environment. An online survey was developed and employed through Google Forms to collect data. The survey was made available to participants on social networks, Facebook@, Whatsapp@, and Instagram@, linked to the research group, during the period 06/15/2020 to 07/15/2020. 1,145 individuals participated in the research. Results: The average age of the participants was 37 years old. They were mostly female, white, Brazilians, with a higher education level, with occupations in the health field, and had religion. They either maintained their family income or suffered a small income decrease. Moreover, they informed that they were caring for social isolation. The most used substances before and after the beginning of the Covid-19 pandemic were alcohol, tobacco, marijuana, although the use of these substances decreased (P< 0.001). Approximately 32% of the participants started using psychoactive substances in this period. Among them, most individuals started by their own initiative. Conclusions: For a better understanding of the pattern use of psychoactive substances during the Covid-19 pandemic and the adverse effects on human behavior and mental disorders, careful longitudinal studies must be developed, due to the great interest in the knowledge of adaptive responses when people’s lives are at risk.

## Introduction

The use of psychoactive substances consists of a global public health problem, which shows high individual and social impact, mostly linked to compromising the mental health of the users and their family members.^1^

Considering the consumption of legal substances, the II National Survey of Alcohol and Drugs (2012) observed a prevalence of adult people (18 years old or older) among the people that have consumed alcoholic beverages in the last 12 months in Brazil. The gender pattern of this indicator consisted of 62% men and 38% women.^2^

Tobacco one of the most determining factors to the Global Disease Burden in the world according to The World Health Organization. Half of the male and one-tenth of the female population, corresponding to 30 million people, will be smokers every year and when they get older. Although the prevalence of smokers has been decreasing among the Brazilians, in 2012 it reached 16.9% of the population, where 21.4% were men and 12.8% were women.^2^

In dealing with illegal substances, Brazil was appointed by the *United Nations Office on Drugs and Crime* as one of the emerging nations where stimulant consumption such as cocaine is increasing, whether in an intranasal form (powder) or smoked form (crack, merla, or oxy). This scenario is against most countries, where the consumption is decreasing.^1,2^

Marijuana is the illicit substance most consumed in the world, and this drug shows the highest prevalence of use in the Brazilian population. Among the adult population, 5.8% affirmed they had used the substance at some time in their lives, which corresponds to 7.8 million of Brazilians. In the last 12 months, 2.5% of Brazilian people declared to have used marijuana, which is equivalent to more than 3 million adults across the country.^2^

Additionally, 3.8% of the adult population affirmed to have used cocaine once in their lifetime, which is equivalent to 5 million of Brazilian people aged 18 and over. The prevalence of cocaine use in the last 12 months among the adult population is 1.7% (more than 2 million Brazilians).^2^

The harmful use and dependence of psychoactive substances, legal or illegal, are associated with physical and mental deterioration. The model of disorders caused by the use of substances is a result of a process where multiple factors interact and influence the behavior of drug use, and the poor judgment to consume a particular substance.^3^

Nowadays, there is sufficient evidence proving that addiction is a mental disease. Changes in the structure and neurochemistry in the brain of drug users affect crucial processes, which implies to transform the voluntary behavior of using drugs into compulsive use. Several factors can interact in the development of drug addiction. The central element is the usage behavior itself. The decision to use a drug is influenced by immediate social and psychological situations, as well as by the remote history of the individual.^3^

Severe Acute Respiratory Syndrome Coronavirus 2 (SARS-CoV-2) is a new respiratory disease initially identified in Wuhan, China, in December 2019, which subsequently spread to many countries around the world.^4,5^ The Covid-19 pandemic has shown itself as a threat to human health, exposing the population to a life condition marked by emotional stress and unclear consequences of mental health.

Covid-19 has a mortality rate around 2%, ^5^ and can be lethal in its initial phase, due to massive alveolar injury and consequent progressive respiratory failure.^4, 6^

The first case of the Covid-19 disease in Brazil was reported in February 2020. ^7^ In March 2020, the Health Ministry declared the community transmission of coronavirus throughout the national territory.^8^

Several protocols were adopted to control and combat the pandemic due to a very fast transmission potential, and the inexistence of effective medications for curative treatment and vaccines to prevent the disease. These protocols include the adoption of social distance, respiratory etiquette through the use of face masks, hand hygiene, and body temperature control.^9^

In this way, the context of the Covid-19 pandemic can be portrayed as a stressful life event, mainly due to the actions that governments worldwide have taken to mitigate the speed of viral spread. Therefore, it carries great potential to cause psychological and emotional damages, to a greater or lesser extent.

The protocols for mitigating the transmission of Covid-19 are embodied in health protection actions aiming to limit the movement of people and agglomerations, varying from social distance to the total blocking or confinement. However, this social limitation is not exempt from consequences, once it can become a trigger to the use of psychoactive substances. The use of these substances may appear as an individual reflex response against the new way of being in the world suddenly imposed by this invisible threat.

The general objective of this work was to investigate the use of psychoactive substances in the context of the Covid-19 pandemic in Brazil. The specific objectives are as follows: a) To know the most used psychoactive substances in the Covid-19 pandemic; b) To examine the frequency of use and the number of doses of psychoactive substances in the context of the Covid-19 pandemic; c) To correlate the frequency of use and the number of doses of psychoactive substances before and after the begging of the pandemic, and; d) To identify the occurrence of the first use of psychoactive substances during the Covid-19 pandemic.

## Methods

A cross-sectional and prospective observational study was undertaken in a virtual environment.

### Sample

A non-probabilistic sampling of voluntary type. Individuals were invited to participate in online research through social media, Facebook@, Whatsapp@, and Instragram@, linked to the research group. The sample answered the online survey using electronic devices such as tablets, smartphones, and computers of all types via internet access. The individuals were only able to answer the online survey upon reading, agreeing, and understanding the Informed Consent Form (ICF), accepting the invitation, and being 18 years old or older.

### Research instrument

The research instrument consisted of an online survey, prepared by the researchers using the Google Forms®, a free search management app. The self-administered online survey was composed of 34 questions and divided into two parts: a general part, with 17 questions regarding sociodemographic and psychosocial characteristics; and a specific part, embracing mental health conditions, behavior associated with the use of psychoactive substances, and possible changes in this behavior, during the Covid-19 pandemic. All answers were anonymous and without any individual identification.

### Data collection

The online survey on the digital platform Google Forms® was made available on the social media, Facebook@, Whatsapp@, and Instagram@, linked to the research group, aiming to invite the public to participate in online research. The research instrument for data collection was maintained with available access 24 hours a day, from 06/15/20 to 07/15/20.

### Ethics Aspects

The present study is in agreement with the recommendations of Resolution 466/2012, which provides ethical aspects in research with human beings in all phases. The research is registered at Plataforma Brasil/CONEP as CAAE 33505120.0.0000.5332. The research was approved according to the substantiated opinion number 4.086.949, by the Research Ethics Committee of São Pedro Psychiatric Hospital, Porto Alegre – RS.

### Statistical analysis

The data collection was made through Google Forms, followed by exportation to Excel, where the collected data was processed. The statistical analysis was carried out using the software IBM SPSS statistics v.20.0. Categorical variables were described by frequency and percentage. Quantitive variables were described by means and standard deviations. The tests Fischer’s Exact or Chi-square with Yates correction were associated with the categorical variables according to the distribution of frequencies in the different categories. Quantitative variables were compared using the t-Student test for independent samples or Analysis of Variance (ANOVA) according to the category’s numbers. Tukey Post-Hoc test was employed aiming at multiple comparations. The categorical variables before and after the Covid-19 pandemic were associated with the McNemar test. The significance level considered for the established comparisons was of 5%.

## Results

Data collection was made from 1156 participants, where 11 were excluded. From those, 6 were younger than 18 years old, and 5 did not accept to participate in the research. The whole sample totalized 1145 participants. The average age of the participants was 37 years old, ranging from 18 to 82 years old. Most of them were female, Brazilian, with high education levels. Moreover, most of them have occupations in the health area, were white, and had religion in which they affirmed to find comfort. In addition, participants with a spouse and cohabitating adults were more frequent. The characteristics of the participants are summarized in Table 1.

**Table 1.** Characteristics of the participants, n=1145.

| **Characteristics** |  | **Descriptive measures** |
| --- | --- | --- |
| Age, average ± SD | | 37.4±1.4 |
| Sex, female n (%) |  | 750 (65.5) |
| Country, Brazil, n (%) |  | 1130 (98.7) |
| Education level*, n (%) |  |  |
|  | Complete or incomplete Elementary School | 14 (1.2) |
|  | Complete or incomplete High School | 77 (6.7) |
|  | Complete or incomplete University Education | 545 (47.7) |
|  | Postgraduate | 506 (44.3) |
| Main practice area, n (%) |  |  |
|  | Health | 537 (46.9) |
|  | Other professions | 211 (18.4) |
|  | Student | 95(8.3) |
|  | Education | 92 (8.0) |
|  | Commerce | 58 (5.1) |
|  | Retired | 49 (4.3) |
|  | Industrial | 39 (3.4) |
|  | Unemployed | 33 (2.9) |
|  | Security | 15(1.3) |
|  | Transport | 13(1.1) |
|  | Cleaning | 3 (0.3) |
| Color or race/ethnicity, n (%) |  |  |
|  | White (European/Western descendants) | 734 (64.1) |
|  | Brown (descendant from individuals with different colors and ethnicities - Miscegenation) | 332 (29.0) |
|  | Black (African/Afro-Brazilian descendants) | 61 (5.3) |
|  | Yellow ( Asian/Eastern descendants) | 15 (1.3) |
| Have religion or spiritual belief, n (%) |  | 961 (83.9) |
| How comfortable is it for you to have a religion? n (%) |  |  |
|  | A lot | 667 (58.3) |
|  | Middling | 257 (22.4) |
|  | Very little | 116 (10.1) |
|  | Nothing | 105 (9.2) |
| Marital status, n (%) |  |  |
|  | Married, living together | 547 (47.8) |
|  | Single | 392 (34.2) |
|  | Married, living alone | 200 (17.5) |
|  | Widower | 6 (0.5) |
| People who they live together, n (%) |  |  |
|  | Living alone | 193 (16.9) |
|  | 1 | 267 (23,3) |
|  | 2 | 286 (25,0) |
|  | 3 | 216 (18,9) |
|  | 4 | 116 (10,1) |
|  | 5 | 48 (4,2) |
|  | More than 5 | 19 (1,7) |

\* 3 participants did not answer. SD: Standard Deviation.

In dealing with the changes in the daily life, most participants try to follow the sanitary recommendations, i.e., to keep the distance from other people, to reduce the human contact, and not to visit elderly people. Most of them are either still working or staying at home and just going out for essential purchases. Regarding the occupation, most of them continued working and studying, either by attending to the workplace or working from home. Most participants either maintained their family income or suffered a small income decrease. Characteristics of the individuals related to the routine, occupation, and income are shown in Table 2.

**Table 2.** Descriptive table about the changes in the routine, occupancy, and income during the
Covid-19 pandemic, n=1145.

| **Characteristics of routine, occupancy, and income** | **n (%)** |
| --- | --- |
| During the Covid-19 pandemic, in which intensity did you or are you doing contact restrictions with the people? |  |
| I tried to take care, to stay away from people, to reduce contact a little, did not visit elderly people, but I kept working. | 517 (45.2) |
| I stayed at home, only going out for essential purchases (pharmacy, supermarket) | 502 (43.8) |
| I stayed strictly at home, only going out for the need for health care | 101 (8.8) |
| I did nothing and lived a normal life | 25 (2.2) |
| How did the Covid-19 pandemic affect your occupancy? |  |
| I kept working/studying | 438 (38.3) |
| I kept working/studying, but at home | 368 (32.1) |
| I did not work before the pandemic, and I continued without work | 181 (15.8) |
| I did not work, but I kept my job | 82 (7.2) |
| I lose my job | 35 (3.1) |
| I got paid vacations | 19 (1.7) |
| I started working after the pandemic | 22 (1.9) |
| How did the Covid-19 pandemic affect your family income? |  |
| It was maintained at the same | 468 (40.9) |
| It decreased a little | 406 (35.5) |
| It suffered a harsh decrease | 202 (17.6) |
| It increased | 47 (4.1) |
| We ran out of income | 22 (1.9) |

When they were asked about their mental health, approximately 37% reported a disorder. Most of them were diagnosed with anxiety, frequently followed by a mood disorder and attention-deficit/hyperactivity disorder. Almost 45% of the participants commonly used some medication and 8% started using some medication after the beginning of the Covid-19 pandemic. Approximately 10% had to increase their medication. Among the participants who had their doses increased, 38% affirmed having done it on their own initiative. These results are summarized in Table 3.

**Table 3.** Descriptive table about the mental health of the participants.

| **Characteristics of mental health** | **n (%)** |
| --- | --- |
| Presence of some disorder, n=1145 | 425 (37.1) |
| Disorder frequency, n=1145 |  |
| Anxiety | 244 (21.3) |
| Mood disorder | 132 (11.5) |
| Attention deficit / hyperactivity disorder | 22 (1.9) |
| Eating disorders | 16 (1.4) |
| Obsessive-compulsive disorder | 7 (0.6) |
| Chemical addiction | 5 (0.4) |
| Schizophrenia and other psychotics disorders | 3 (0.3) |
| *Borderline* | 1 (0.1) |
| Current use of some medication, n=1145 | 512 (44.7) |
| Started to use some medication after the beginning of the pandemic, n=991 | 80 (8.1) |
| Need to increase the medication, n=969 | 99 (10.2) |
| If increased doses, reason n=156 |  |
| By their own initiative | 59 (37.8) |
| By medical advice | 97 (62.2) |

Regarding the frequency of substance usage before the pandemic, alcoholic beverages were the most frequent, followed by tobacco, marijuana, and psychotropic medications. After the Covid-19 pandemic started, these substances were kept as most used, in the same order. A significant reduction in the consumption of alcoholic beverages, tobacco, and marijuana occurred (P<0.001 for all of them). The use of psychotropic medications, cocaine, analgesics, and other mentioned substances did not change significantly. LSD, MDMA, or MD suffered a reduction in their use, statistically significant (P=0.008). When the use of any substance was evaluated, there was a significant reduction before and after the pandemic, where 12.3% of the participants stopped using some substance during the pandemic, and 2% started (from 59.2% who used it before to 48.9% who used it after; P< 0.001). The descriptive and comparative data about the use of psychoactive substances before and after the beginning of the Covid-19 pandemic are presented in Table 4.

**Table 4.** Comparative and descriptive table about the use of psychoactive substances before and
after the start of the Covid-19 pandemic. n =1145.

| **Substances** | **Before** | **After** | **Stopped** | **Started** | **P** |
| --- | --- | --- | --- | --- | --- |
| Alcoholic beverage | 570 (49.8) | 439 (38.3) | 146 (12.8) | 15 (1.3) | **<0.001** |
| Tobacco | 174 (15.2) | 134 (11.7) | 46 (4.0) | 6 (0.5) | **<0.001** |
| Marijuana | 117 (10.2) | 91 (7.9) | 30 (2.6) | 4 (0.3) | **<0.001** |
| Psychotropic medication | 100 (8.7) | 89 (7.8) | 29 (2.5) | 18 (1.6) | 0.144 |
| Cocaine | 10 (0.9) | 8 (0.7) | 3 (0.3) | 1 (0.1) | 0.625 |
| LSD/MDMA/MD | 9(0.8) | 1 (0.1) | 8 (0.7) | - | **0.008** |
| Analgesic and other mentioned substances | 4 (0.3) | 1 (0.1) | 3 (0.3) | - | 0.250 |
| Some substance | 678 (59.2) | 560 (48.9) | 141 (12.3) | 23 (2.0) | **<0.001** |

Data presented by n (%) and associated with the McNemar test, that compare data before and after some event.

When the frequency of use and quantity of doses of psychoactive substances were evaluated, a significant change in the dose and frequency before and after the Covid-19 pandemic was noted. There was an increase in the number of subjects who do not use any substance However, among the people who used to consume some substance, there was an increase in those who have used more than 3 times a week. Regarding the doses among the people who have consumed some substance, there was an increase in those who consume 6 or more doses. These data are exhibited in Table 5.

**Table 5.** Descriptive and comparative table of frequency and dose of psychoactive substances
before and after the beginning of the Covid-19 pandemic. n =1145.

| Frequency and dose of psychoactive substances |  | Before | After | P |
| --- | --- | --- | --- | --- |
| Frequency |  | n=1145 | n=1145 | <0.001 |
|  | Do not use any substance | 454 (39.7) | 565 (49.3) |  |
|  | Lesser than 1 day per week | 188 (16.4) | 117 (10.2) |  |
|  | 1-2 days per week | 261 (22.8) | 154 (13.4) |  |
|  | 3-4 | 103 (9.0) | 122 (10.7) |  |
|  | 5-6 | 26 (2.3) | 67 (5.9) |  |
|  | Daily | 113 (9.9) | 120 (10.5) |  |
| Dose |  | n=610 | n=542 | 0.006 |
|  | 1 | 170 (27.9) | 149 (27.5) |  |
|  | 2 | 150 (24.6) | 117 (21.6) |  |
|  | 3 | 92 (15.1) | 88 (16.2) |  |
|  | 4 | 52 (8.5) | 44 (8.1) |  |
|  | 5 | 37 (6.1) | 29 (5.4) |  |
|  | 6 or more | 109 (17.9) | 115 (21.2) |  |

The data was presented by n (%) and associated with the McNemar test.

Among the 1145 research participants, 32% referred starting the use of psychoactive substances during the pandemic, where 31 (2.7%) did not use any substance before the pandemic, and 335 (29.3%) added some substance to their initial consumption. Most of them started consuming some substance on their own initiative. Approximately 4% of them had to replace the substance, mostly due to the difficulty in access. Most of the participants do not think that the consumption of the substances is beyond their control. Moreover, they do not think it is difficult to be without any of these substances, they are not worried and have no desire to stop. The description of the occurrence of the first use of psychoactive substances in the context of the Covid-19 pandemic, as well as consumer perception, are presented in Table 6.

**Table 6.** Description of the occurrence of the first use of psychoactive substances in the context of the Covid-19 pandemic, and the consumer perception.

| **Use of psychoactive substances** | **n (%)** |
| --- | --- |
| Started the consumption during the pandemic. n=1145 | 366 (32.0) |
| Started the consumption during the pandemic (did not use to consume anything). n=1145 | 31 (2.7) |
| Added some substance to their initial consumption. n=1145 | 335 (29.3) |
| Reason. n=366 |  |
| On their own initiative | 327 (89.3) |
| By suggestion of other people | 39 (10.7) |
| Need to replace. n=1145 | 49 (4.3) |
| Reason. n=49 |  |
| Cost | 23 (46.9) |
| Access | 26 (53.1) |
| The use of some substances is beyond their control. n=739 |  |
| Always/Almost always | 15 (2.0) |
| Frequently | 33 (4.5) |
| Sometimes | 131 (17.7) |
| Never/Rarely | 560 (75.8) |
| How difficult is it to be without some of these substances? n=762 |  |
| Impossible | 12 (1.6) |
| Very hard | 93 (12.2) |
| A little hard | 204 (26.8) |
| Nothing difficult | 453 (59.4) |
| The fact that you consume some substance worries you? n=711 |  |
| Always/Almost always | 30 (4.2) |
| Frequently | 45 (6.3) |
| Sometimes | 207 (29.1) |
| Never/Rarely | 429 (60.3) |
| Do you want to stop using any of these substances? n=698 |  |
| Always/Almost always | 45 (6.4) |
| Frequently | 66 (9.5) |
| Sometimes | 222 (31.8) |
| Never/Rarely | 365 (52.3) |

## Discussion

The present study was elapsed in Brazil during the Covid-19 pandemic, whose social and economic impacts are shown to compromise the quality of people’s mental health. This occurs mostly because of the strategies adopted to reduce viral transmission, increase the installed capacity, and the qualification of the health system to treat the patients. Anxiety disorders, affective disorders, and other psychological suffering have already been reported as induced by the Covid-19 pandemic. ^10,11^

In this research, most of the participants (59.2%) reported using some psychoactive substances before the pandemic, being the legal most consumed alcohol (49.8%) and tobacco (15.2%), and the illegal, marijuana (10.2%). These results find support in the literature about the prevalence of substance use among the population. ^1,2,3^

The comparison between alcohol, tobacco, and marijuana consumption before and after the Covid-19 pandemic, considering the sample profile, indicated a statistically significant reduction in their consumption. This result goes in the opposite direction of the evidence that has been published by the academic community.^12,13,14,15,16,17^ A decrease in alcohol consumption was also found in a published study, reinforcing a possible change in drinking habits due to the pandemic.^17^ The interpretation of this phenomenon involves several factors, which will be explored here.

In a scenario of recreational use of these substances, the first factor could be focused on the strategies to mitigate the viral transmission of Covid-19. The restriction protocols imposed on shops, bars, restaurants, clubs, and counterparts^18^ and the effective suspension of these places could justify this harsh decrease in alcohol consumption.^19^

However, this topic is controversial because we may be facing a readjustment process, which changes the way of access and utilization of these psychoactive substances, with new social purchasing practices reported by the increase in the sales^16^ in supermarkets, shops, and on the internet,^20^ and the consumption outside the commercial places.

A second factor that could justify the decrease in the consumption of these three identified substances would be the hypothesis of consumption place relocation during the pandemic. People who used to make recreational use of substances in the context of social interaction are now deprived of the social environment to drink and smoke in the company of their social groups. This fact is attributed to the restrictions imposed by social distancing and supported by the closure of entertainment venues. In this way, the deprivation of social contact in locations and situations associated with the entertainment of social groups would be a limiting factor to the consumption of these substances.

In the opposite direction, the increased consumption of psychoactive substances reported in other publications could be supported by the innovative practices of purchase and consumption in the context of the Covid-19 pandemic. These practices were symbolized by a new trend marked by the reinvention of entertainment ways, such as virtual events with real-time interaction and online appeals to consumption.

Although there was a reduction in the consumption of psychoactive substances in the studied sample, it is interesting to observe the results regarding the frequency of use and consumption dosage. The frequency of use of psychoactive substances by people who used to consume substances more than 3 days a week increased after the Covid-19 pandemic. Moreover, the number of daily doses consumed by people who used to consume 6 or more doses increased.

These findings encourage the reflection about the possibility of addictive behavior in these cases because most of the sample reported that the use of some substance is not beyond their control. Furthermore, most of them reported they were neither feeling difficulty to be without any of these substances nor being worried or wish to stop consuming them.

It is important to emphasize the fact of 32% of the analyzed sample started consuming some substance during the Covid-19 pandemic, by their own initiative. The meaning attributed to the expression “started using some substance” deserves to be highlighted. It means, on the one hand, that the individual may have added a new substance to others already in use, which occurred in 29.3% of the sample. On the other hand, it means that individuals who did not use to consume some substance before the pandemic and started using during this period, which was reported by 2.7% of the sample.

Thus, among the individuals who reported using some substance before the pandemic, part of them increased the frequency – days of consumption - and the number of doses used during the pandemic. Another fraction of the sample started the consumption of some substance after the beginning of the Covid-19 pandemic. This part corresponds either to people who started using something that did not use before the pandemic, or who added some substance to others already commonly used.

It is known that the Covid-19 may not directly change the consumption habits of psychoactive substances, but the stress caused by the threat of its infection can increase negative emotions such as anxiety and depression. ^11^ In this sense, the protocols of social distance, from the extremes of social distance to lockdown, reduced the social contact of a large number of people, which affects many aspects of people’s lives and might trigger psychological suffering. This may be responsible for compromising psychosocial functioning, such as occurs in panic disorder, anxiety, and depression.

Scientists and institutions have warned that the Covid-19 pandemic may have increased the risk of substance abuse that causes addiction as well as addiction-related behavior. ^21,22^

## Study limitations

The first limitation attributed to this study is related to the chosen sample, which was made by convenience. The non-probabilistic sample enabled a high response rate. However, the generalization of the results to the Brazilian population, in general, is not possible. External validity is limited to samples of the same profile.

The inclusive criteria for people aged 18 or over have excluded the teenagers of the population, whose marijuana consumption may be higher, and whose alcohol consumption shows singular characteristics, related to parties and groups. This population portion may have changed the consumption behavior during the pandemic, but these changes were not ensured by this research. This age range may be addressed in future studies.

This research may be influenced by memory bias, once the participants were requested to recall habits and behaviors to answer determined questions in the survey.

Finally, the need to send a concise survey that is simple for the participant to complete led to the joint exploration of the psychoactive substances without any distinction between different types of substances. In other words, there were no questions that specified psychoactive drugs one by one and their respective frequencies of use and doses of consumption, before and after the start of the pandemic. Thus, the richness of the data related to the high increase or decrease in the consumption of some substances may have been missed, because the analyses of all substances were addressed as a whole at the same level. Alternatively, two variables were created, called “dose change” and “start of consumption”, which summarized the information about the use of psychoactive substances.

## Conclusions

The results obtained by this research revealed a behavior change associated with the use of psychoactive substances in a time of social distance in several degrees of isolation, which has been affecting many aspects of people’s life.

Nonetheless, careful longitudinal studies must be developed for a better understanding of the use pattern of psychoactive substances in the context of the Covid-19 pandemic. They would allow understanding the adverse effects on behavior and other mental disorders linked to the use of psychoactive substances during the pandemic. Furthermore, these studies would contribute to investigating people’s adaptive responses at a time when their lives are at risk.

## Data Availability

All necessary patient/participant consent has been obtained and the appropriate institutional forms have been archived.

## Notes

### Competing Interest Statement

The authors have declared no competing interest.

### Funding Statement

No funding was received for this study.

### Author Declarations

The present study is in agreement with the recommendations of Resolution 466/2012, which provides ethical aspects in research with human beings in all phases. The research is registered at Plataforma Brasil/CONEP as CAAE 33505120.0.0000.5332. The research was approved according to the substantiated opinion number 4.086.949, by the Research Ethics Committee of Sao Pedro Psychiatric Hospital, Porto Alegre, RS.

